# Metastatic spine disease alters spinal load-to-strength ratios in patients compared to healthy individuals

**DOI:** 10.1101/2025.01.06.25320075

**Authors:** Dennis E. Anderson, Mario Keko, Joanna James, Brett T. Allaire, David Kozono, Patrick F Doyle, Heejoo Kang, Sarah Caplan, Tracy Balboni, Alexander Spektor, Mai Anh Huynh, David B. Hackney, Ron N. Alkalay

**Author notes:** Address correspondence to: Ron N. Alkalay, PhD Associate Professor in Orthopedic Surgery Center for Advanced Orthopedic Studies Beth Israel Deaconess Medical Center, 1 Overland Street, Boston, MA, 02215, USA.

## Abstract

Pathologic vertebral fractures (PVF) are common and serious complications in patients with metastatic lesions affecting the spine. Accurate assessment of cancer patients’ PVF risk is an unmet clinical need. Load-to-strength ratios (LSRs) evaluated in vivo by estimating vertebral loading from biomechanical modeling and strength from computed tomography imaging (CT) have been associated with osteoporotic vertebral fractures in older adults.

Here, for the first time, we investigate LSRs of thoracic and lumbar vertebrae of 135 spine metastases patients compared to LSRs of 246 healthy adults, comparable by age and sex, from the Framingham Heart Study under four loading tasks. Findings include: (1) Osteolytic vertebrae have higher LSRs than osteosclerotic and mixed vertebrae; (2). In patients’ vertebrae without CT observed metastases, LSRs were greater than healthy controls. (3) LSRs depend on the spinal region (Thoracic, Thoracolumbar, Lumbar). These findings suggest that LSRs may contribute to identifying patients at risk of incident PVF in metastatic spine disease patients. The lesion-mediated difference suggests that risk thresholds should be established based on spinal region, simulated task, and metastatic lesion type.

---

In 2022, an estimated 1.9 million new cancer cases occurred in the U.S ^1^. Vertebral bone metastases (BM) are common in patients with cancer ^2^, affecting up to 70% of patients with advanced cancer ^3^. Radiologically, bone metastases appear as osteosclerotic (bone-forming), osteolytic (bone-destroying), or a combination of both, termed mixed lesions ^4^. At the bone tissue, BM disrupts the bone’s microarchitecture ^5,6^, tissue properties ^7,8^, composition ^9^ and cellular homeostasis ^10^, causing the degradation of the bone’s mechanical properties ^8,11^. Pathologic vertebral fractures (PVF), a significant clinical complication affecting up to 39% of cancer patients treated with radiotherapy for spinal BM ^12^, cause severe impairment of patient’s quality of life ^13^, higher health costs ^14^, may lead to neurological deficits due to metastatic epidural spinal cord compression (MESCC) affecting to 15-20% of patients with metastatic spine disease ^15^, and shorten patient survival ^16–18^ and 3-year life expectancy ^13,16^. Current clinical diagnostic protocols have low to, at best, moderate sensitivity and specificity for predicting PVF ^19–21^. Establishing factors to allow precise individualized prediction of PVF risk before catastrophic pain or neurologic deficits occur remains a critical, unmet clinical need for managing patients with spinal metastatic disease.

From a mechanical perspective, PVF is caused when the metastatic bone lesion has degraded the vertebra’s strength such that it can no longer sustain loads of daily living. Conceptually, PVF may occur if the value of the applied loading to the lesioned vertebrae exceeds its strength, i.e., the Load-Strength Ratio (LSR) is greater than one ^22^. Vertebral loading, produced largely via the activity of the trunk and abdominal muscles ^23^, varies greatly according to the individual’s body weight ^24^, spine curvature ^25–27^, and vertebral level ^26^. This activity is required to balance the applied external loads generated by a specific task, affects the motion required by this task ^28^, and provides mechanical stability to maintain spinal posture ^29,30^. Compared to older subjects without prevalent osteoporotic vertebral fractures (OVF), age and sex-matched subjects with OVF showed lower vertebral strength ^31–34^ and higher LSRs in the thoracolumbar spine region, regions where VF occurs most frequently, in response to flexed or upright postures carrying weights ^26,35^. These findings suggest that LSR value and pattern are important considerations in osteoporosis-related vertebral fracture risk. Although the role of spinal loading as a contributor to PVF risk has not been similarly studied, our recent study, evaluating 11 cadaveric spines from patients with bone metastases, found bone metastasis to modify LSR at the vertebral level. Vertebral levels containing osteosclerotic or mixed lesions were estimated to have lower LSRs, and conversely, osteolytic vertebrae had higher LSRs than healthy norms ^36^. Uniquely, cancer donor vertebrae without radiographic evidence of bone metastasis demonstrated higher LSR than healthy normative values ^36^. These findings indicate that cancer has a systematic effect on the biomechanical properties of the spinal column and suggest that biomechanical analysis to drive LSR may highlight vertebral levels at increased PVF risk in patients, even at levels without CT-identified lesions, the current clinical gold standard for evaluating vertebrae at PVF risk. With metastases and associated PVF occurring throughout the spine, understating the role of patient- and task-specific LSR on the pathologic vertebra is crucial for developing a more comprehensible assessment of fracture risk in patients with metastatic spine disease and determining its potential clinical utility.

This study aims to evaluate the effect of the clinical classification of metastatic bone lesions on task-specific vertebral LSRs in a cohort of cancer patients treated with radiotherapy for spine metastatic disease. Specifically, we aim to 1) evaluate LSRs for key functional activities *in vivo* and compare between patients with metastatic disease and norms in a healthy population; 2) examine whether LSRs from patients with metastatic disease in levels <u>without</u> CT-observable lesions (No Observed Lesion – NOL) differ from norms in a healthy population; and 3) determine whether LSRs differ with metastatic lesion classification (NOL, Osteolytic, Osteosclerotic, and Mixed lesions) in patients with metastatic spine disease. We hypothesized that LSRs will differ between patients with metastatic spine disease and healthy controls, including at levels without CT- observable lesions (NOL), and further that vertebrae classified with Osteosclerotic lesions and Mixed lesions will have lower LSRs than vertebrae with Osteolytic lesions and NOL.

## Results

### Characteristics of spine metastasis patients and normative dataset subjects

The sample included 135 patients with spine metastatic disease, compared to 246 participants from the Framingham Heart Study (FHS) cohort. Table 1 shows the characteristics of demographics and anthropometric measures across different groups. The average age of the study participants is around 64 years old for both groups, with no significant differences (p=0.965). The proportion of men is significantly higher (p=0.001) in the patient cohort (66%) compared to FHS (50%). Accordingly, patients are taller (1.71 *m* vs 1.67 *m*; p<0.001), but there are no differences in the weight of the participants overall (p=0.715). The most common primary cancers were prostate (n = 56), lung (n = 23), renal (n = 14), and breast (n = 11).

**Table 1:**
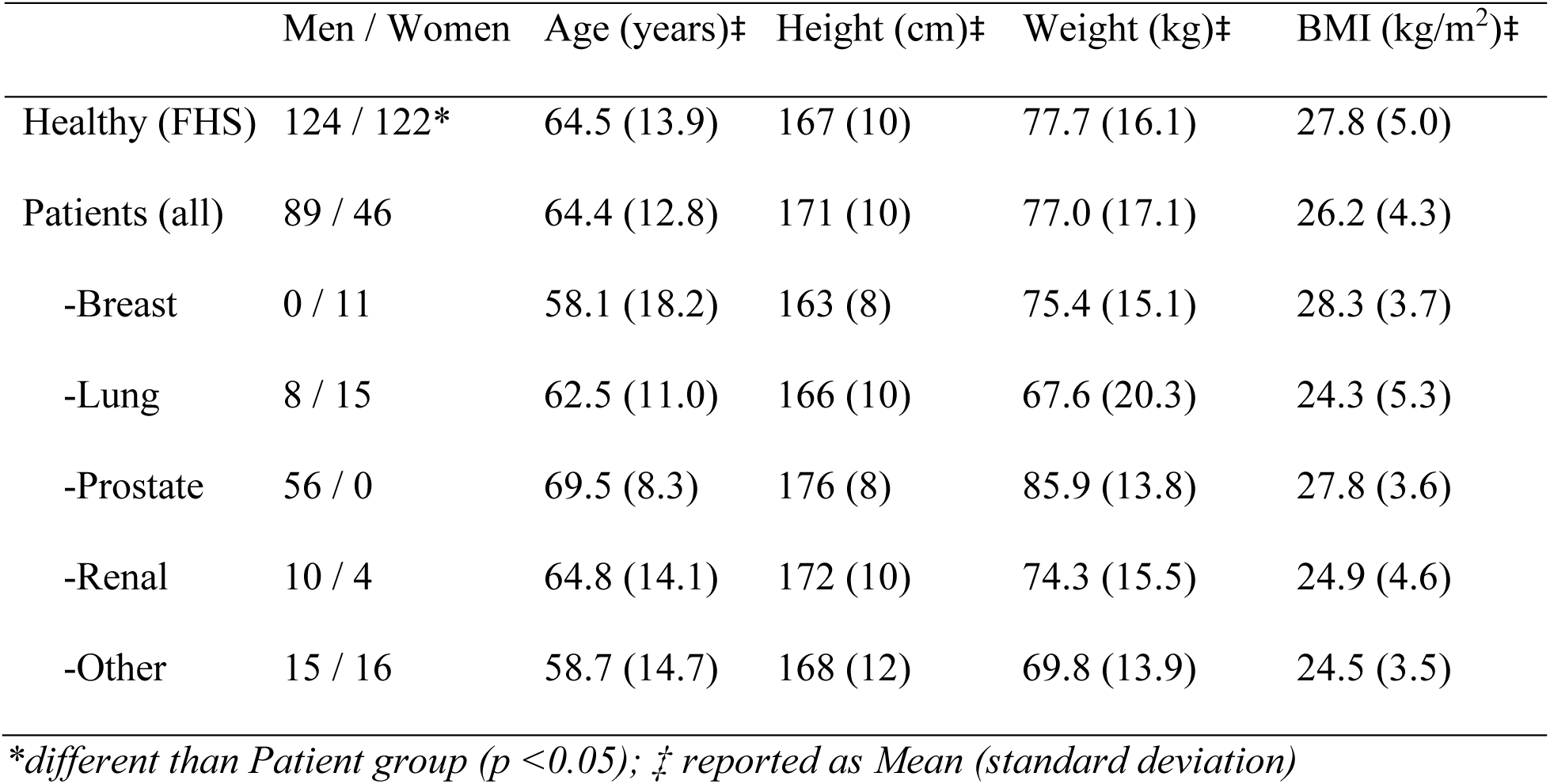
Characteristics of Healthy (FHS) and Patient groups and patients by primary cancer, including counts by sex, age, height, weight, and BMI.

### LSRs for spine metastasis patients and normative dataset subjects

Spine LSRs were evaluated for 1,508 vertebral levels (T4 – L4) in patients and 2,990 in FHS subjects. Within the patient cohort, radiographic review classified 175 (12%) of the vertebrae as osteosclerotic, 84 (6%) as mixed lesion type, 119 (8%) as osteolytic and 1,119 levels (75%) as levels with no lesion observed on the clinical CT. Table 2 shows LSR level counts for each spinal region by study group and lesion type (for patients).

i. <u>Comparison of cancer cohort and FHS subjects</u>: The unadjusted LSRs (Figure 1) are higher in the FHS cohort than spine metastases patients for all tasks (p<0.001), specifically 0.064±0.023 vs. 0.060±0.024 for Neutral Standing; 0.167±0.067 vs. 0.152±0.060 for Standing + Weight; 0.179±0.071 vs. 0.159±0.065 for Flexion + Weight; and 0.107±0.036 vs. 0.093±0.033 for Lateral Bend + Weight. The statistical analyses showed that standardized LSRs were affected by group, age, and spinal region for men and women across all four tasks (Figure 2; Supplemental Tables 1A and 1B). The effects of grouping on standardized LSR showed similar overall trends by region and activity but with some notable contrasts in the findings by sex (Figure 2).
ii. <u>Spine regions demonstrate significant differences in LSR</u>: In men, NOL vertebrae had significantly higher LSR values in the Thoracolumbar and Lumbar regions for the simulated NS than FHS subjects (Figure 2). We found no statistically significant difference when comparing region-based differences in LSR for the remaining simulated tasks (Figure 2). In women, NOL vertebrae had significantly lower LSRs than healthy (FHS) in the Thoracic (NS, S+W, & LB+W), Thoracolumbar (S+W, F+W, & LB+W), and Lumbar (F+W) regions (Figure 2).

**Figure 1:**
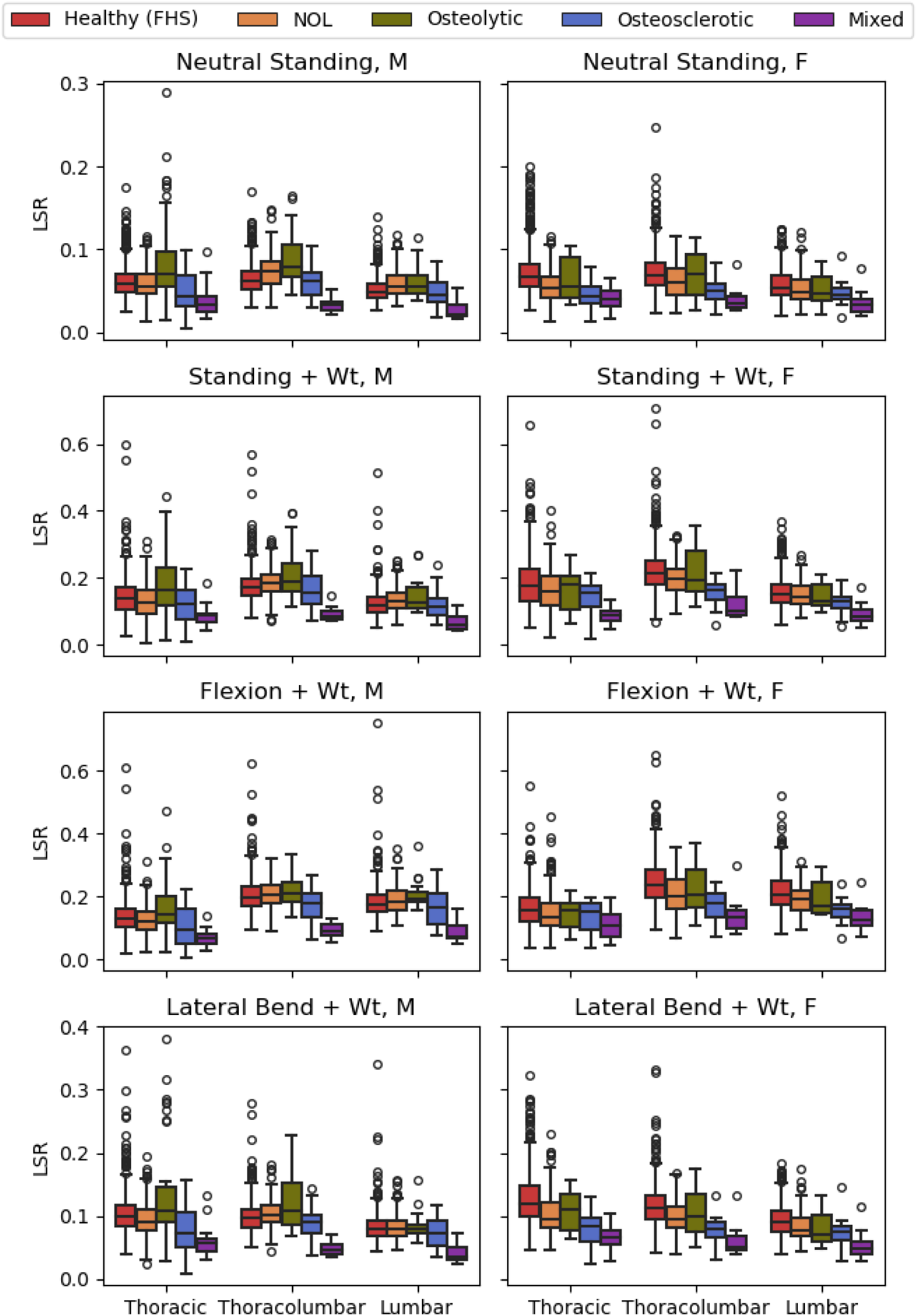
Spine LSRs in healthy people from the Framingham Heart Study (FHS) and patients with spine metastases – grouped by lesion type in three spinal regions – separately for four loading tasks (Rows) and by sex (Columns). NOL = no observed lesion.

**Figure 2:**
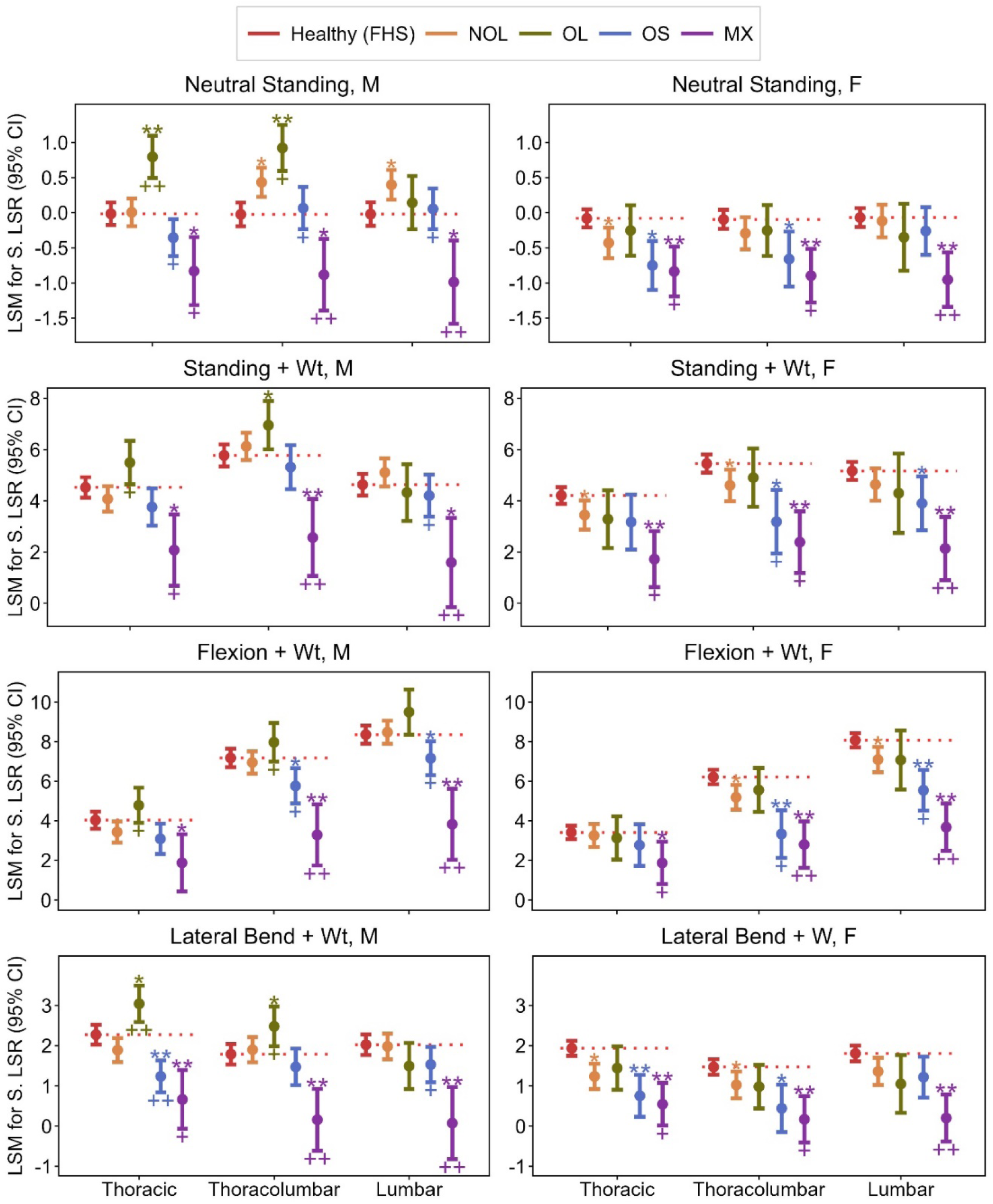
Least Square Mean (LSM) differences in Standardized Spine LSRs (calculated as linear combinations from the mixed effects regression model, normalized to the Neutral Standing task) comparing patients with spine metastases and healthy people from the Framingham Heart Study (FHS), grouped by lesion type in three spinal regions – separately for four loading tasks (Rows) and by sex (Columns). NOL = no observed lesion; OL = osteolytic; OS = osteosclerotic; MX = mixed. *P < 0.05 & **P < 0.001, for comparison vs. FHS. ^+^P<0.05 & ^++^P<0.001 for comparison of lesions vs. NOL; P values are adjusted for multiple testing by the FDR correction

**Table 2:** Counts of vertebral levels included in the analysis for each spinal region by study group and lesion type (for patients).

|  | Thoracic | Thoracolumbar | Lumbar |
| --- | --- | --- | --- |
|  | (T4 – T10) | (T11 – L1) | (L2 – L4) |
| Healthy (FHS) | 1,614 | 662 | 714 |
| Patients (all types) | 789 | 386 | 333 |
| -No Lesion Observed | 607 | 284 | 239 |
| -Osteolytic | 58 | 37 | 24 |
| -Osteosclerotic | 81 | 43 | 51 |
| -Mixed | 43 | 22 | 19 |

In men, Osteolytic levels showed higher LSRs than healthy (FHS) levels in the Thoracic (NS & LB+W) and Thoracolumbar (NS, S+W, & LB+W) regions. In contrast, LSRs for Osteolytic levels in women were comparable or lower than healthy levels (FHS) for all levels and activities, and none of these differences were statistically significant. Osteosclerotic levels in men had lower LSRs than healthy (FHS) levels in the Thoracic region (LB+W only) and the Thoracolumbar and Lumbar regions (F+W only). In women, osteosclerotic levels had lower LSRs than healthy (FHS) levels in the Thoracic (NS & LB+W), Thoracolumbar (all), and Lumbar (S+W & F+W) regions. Mixed levels had lower LSRs than healthy (FHS) levels in both men and women, in all spine regions, and in all activities.

iii. <u>Bone metastasis type significantly affects LSR in cancer patients</u>: Osteolytic levels in men had higher LSRs than NOL vertebrae for Thoracic (all) and Thoracolumbar (NS, F+W, & LB+W) regions. In contrast, for all activities and spinal regions, osteolytic levels LSRs were either comparable or lower than NOL vertebrae LSRs in women, the differences not statistically significant. In Men, Osteosclerotic levels had lower LSRs than NOL vertebrae for the Thoracic (NS & LB+W), Thoracolumbar (NS & F+W), and Lumbar (all) regions. In women, osteosclerotic levels had lower LSRs than in the NOL vertebrae for the Thoracolumbar (S+W & F+W) and Lumbar regions (F+W only). Finally, Mixed levels had lower LSRs than NOL vertebrae in both men and women at all spine regions and for all activities (but not reaching significance for men in the Thoracic region for F+W simulated activity).

## Discussion

In cancer patients with spinal bone metastases, the risk of PVF is a critical factor in management ^3,37^. However, the biomechanics of the PVF risk are poorly understood. Understanding the influence of biomechanical factors and the effect of daily activities on these factors as contributors to this risk may allow progress toward precise individualized PVF risk prediction in patients with spinal bone metastases, a significant medical need. This study applied spinal musculoskeletal models derived from clinical CT data of cancer patients with spinal bone metastases, and of a healthy cohort of subjects of a similar age and sex distribution as a comparison, to estimate individualized vertebral- specific loading and strength values in response to four common daily activities. We used the biomechanical metric of the load-to-strength ratio to identify lesion- and task-specific differences between the cancer and healthy cohorts. For the first time, our study demonstrates biomechanical differences in LSR in patients with spinal bone metastases compared to healthy populations. We further observed the effect to be dependent on clinical lesion classification among patients, with osteosclerotic and mixed vertebrae having lower LSRs than those in healthy subjects. We found osteolytic vertebrae to have higher LSRs in men, but surprisingly, in women, we found LSRs for osteolytic vertebrae were not significantly different than healthy subjects. Uniquely, and particularly in women, we observed that patients’ vertebrae with no CT evidence of bone lesion (NOL) had LSRs lower than the reference non-cancer cohort when adjusted for age and stratified by sex. This finding suggests that cancer and its associated treatments have a systemic effect on the patient’s musculoskeletal spine system, and thus, such vertebrae may not be considered “healthy,” affecting their risk of fracture. Our study proposes that clinical evaluation of LSRs may provide an important metric to the assessment of the effect of cancer patients’ daily activities on the capacity of the lesioned vertebrae to carry the imposed spinal loading, likely a key determinant of fracture risk, with important implications for patient management and risk reduction.

Previously, we have shown that the CT-based estimate of vertebral compressive strength used here is well-correlated with measured strength in metastatic vertebrae ^36^ and that lower LSRs in levels with mixed and osteosclerotic lesions were related to higher estimated strengths in those vertebrae^36^. We note that this was a cadaveric study limited to specimens from 11 individuals, 3 women and 8 men, while the current analysis examined 135 patients, 46 women and 89 men; our findings for the clinical cohort show similar trends for strength (Figure 3) and estimated LSRs, particularly the reduced LSRs associated with osteosclerotic and mixed lesion types ^36^, providing much stronger evidence. The prior study also indicated higher LSRs related to osteolytic and NOL classifications, but this was less consistent across activities in the present study. Here, we found this trend within the patients but not in comparison to the healthy cohort. Specifically, although male patients with osteolytic vertebrae showed an inconsistent trend for LSRSs in the lumbar region, the thoracic and thoracolumbar osteolytic vertebrae had higher LSRs than the healthy male cohort, suggesting a higher risk of PVF in osteolytic vs osteosclerotic vertebrae. This finding is consistent with clinical observations ^37^. Unexpectedly, in female patients, osteolytic vertebrae had comparable LSRs to healthy subjects independent of region or activity simulated. This finding is inconsistent with clinical experience of high risk of PVF in osteolytic vertebrae ^37^. Examination of LSRs by primary cancer suggests female breast and renal cancer patients had higher LSRs than FHS subjects (Figure 4), indicating a higher risk of PVF, a finding consistent with clinical experience ^37^. By contrast, we found that lung and other cancer patients had lower LSRs (Figure 4). We trace this difference to both groups’ markedly lowering musculoskeletal model-predicted vertebral compressive load compared to FHS subjects. For example, in female lung patients, the estimated vertebral compressive load was markedly lower than the FHS controls, with a mean difference of -11%, while differing by 0.5% difference in estimated strength, yielding lower LSRs. With these two groups being our study’s largest groups of female patients, the overall finding resulted in an overall lack of difference for LSRs in female patients with osteolytic lesions despite the clear differences among cancer patients (Figure 4). While examination by primary cancer was not a stated goal of the current study, this provides motivation and support for future efforts to consider variations in LSR with primary cancer as well.

**Figure 3:**
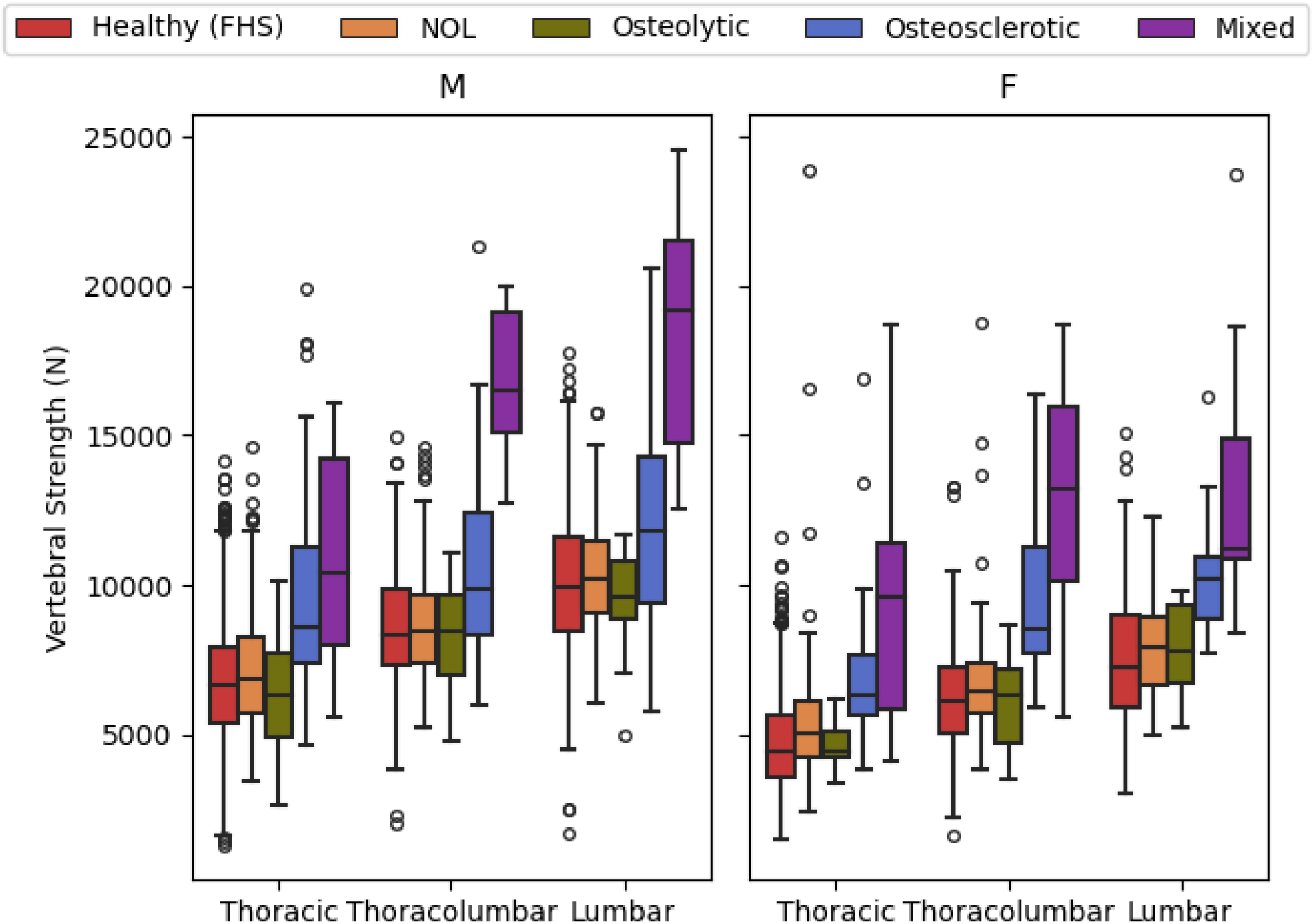
Compressive vertebral strength estimated from CT data in healthy people from the Framingham Heart Study (FHS) and patients with spine metastases – grouped by lesion type and spinal region. NOL = no observed lesion.

**Figure 4.**
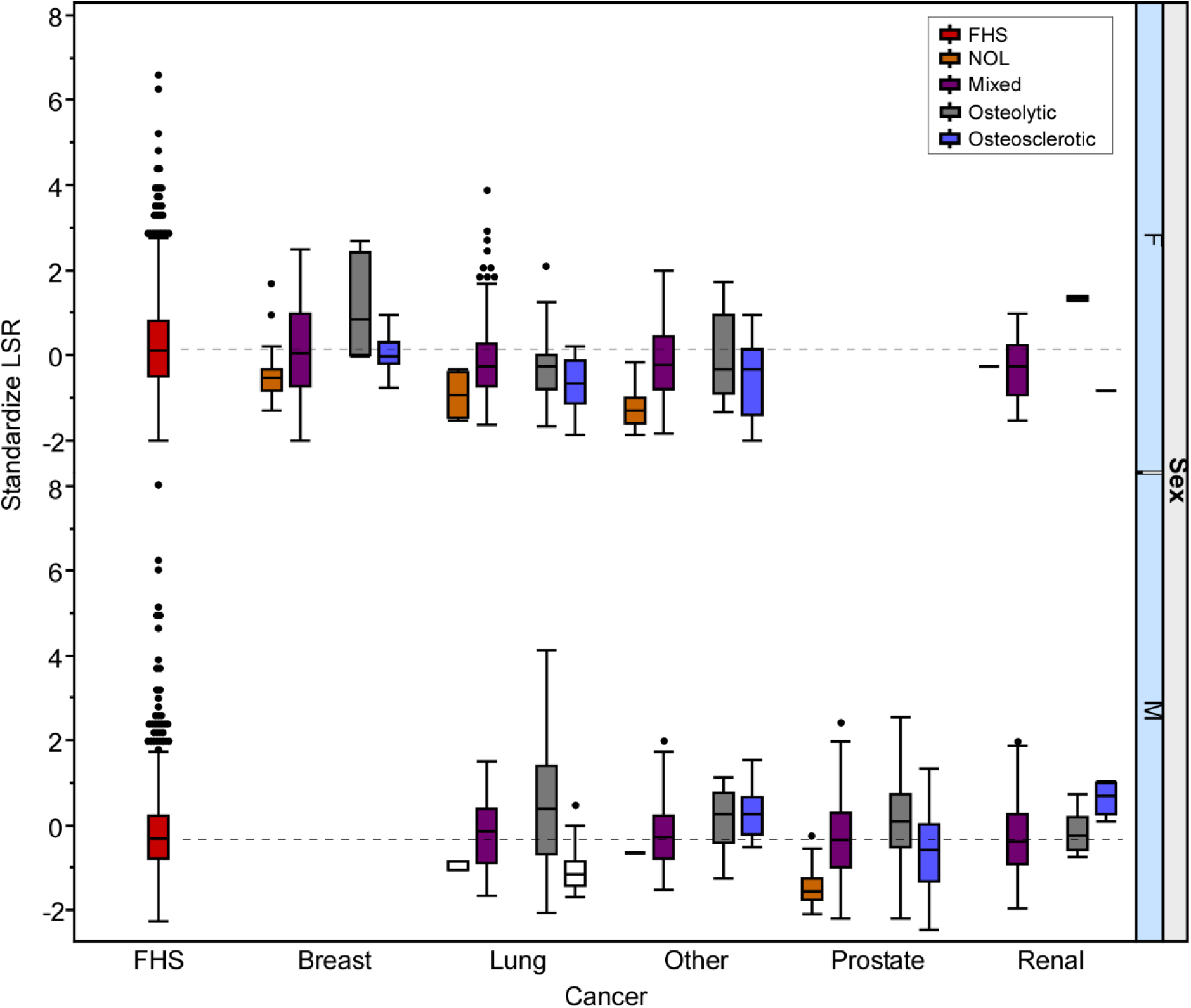
Standardized load to strength ratio (LSR) estimated from clinical CT derived musculoskeletal models in patients with spine metastases and in healthy people from the Framingham Heart Study (FHS)- grouped by primary cancer and sex. NOL = no observed lesion. Dotted line indicates mean LSR value for FHS subjects.

The spine forms a complex multi-articular system in which, under the nervous system’s control, the muscles actively confer mechanical stability, actuation and motion required for performing daily tasks ^38^. Spinal instability, which often occurs with neoplastic spinal disease ^39^, has largely been attributed solely to pathologic osseous changes that compromise the vertebral mechanical integrity, resulting in an increased risk of vertebral fracture ^40^. However, there remain uncertainties related to the association between cancer, bone metastases, and, ultimately, the mechanisms of failure leading to vertebral fracture ^41,42^, which is rarely a simple compression fracture. Paraspinal muscles play a critical role in the stabilization and mobility of the spine ^43^, with paraspinal muscles’ lower cross-sectional area (CSA) reported to be associated with low back pain ^44,45^ and disability ^46^. Cancer patients routinely present myopathic changes ^47^ with Sarcopenia, broadly characterized as the loss of muscle mass, strength, and functional decline, a common sequela of cancer ^48,49^. The etiology of Sarcopenia in cancer is multifactorial, with overlapping causes such as the pathologic process of cancer itself, side effects from cancer treatments, and advanced age.

Meanwhile, model-predicted spine loading is sensitive to individualized muscle parameters (size and location) and spinal posture in complex ways, which can alter loading estimates by up to 54% ^27^. Muscle and spine posture parameters may not be normal in patients with cancer, but whether and how these vary with primary cancer and treatment has not been carefully investigated. These factors could affect musculoskeletal loading in cancer patients, manifesting as differences in LSRs. Furthermore, Radiation greatly affects muscular tissue through fibrosis, weakness, fatigue, and altered neural control ^50^. Moreover, fatigue and weakness are documented side effects in patients receiving radiation ^51^. Thus, while data on the specific effects of irradiation on *in vivo* spine muscle function in patients with spinal metastatic disease are lacking, it is plausible that radiation therapy exacerbates the risk of PVF through neuromuscular changes that affect spine loading and stability. This uncertainty may explain our finding that LSRs do not increase uniformly in metastatic spines compared to healthy controls, with complex variations apparently related to sex, region, task, lesion type, and primary cancer.

The study has several noted limitations. Our patients were recruited from patients treated with radiotherapy, leading to a cohort of patients with multiple primary cancers. In this study, we chose to report patient characteristics across the most common primary types. However, due to the resulting patient number per primary, we lacked the power to examine how primary affects LSR outcomes reliably. This remains a limitation, and future efforts may address whether primary cancer modifies LSR. However, the study findings related to the effect of lesion type on LSR are of clinical interest nonetheless. The study groups of patients and the healthy cohort were not explicitly matched by age, sex, height and weight. However, the healthy subjects’ mean and range of ages closely matched that of our patient group. While not particularly different otherwise, the patient group was predominantly male. Our analyses were stratified by sex, making this a minor limitation. We performed analyses to adjust for height, weight and BMI, but this had little effect and was not included in the reported results.

Our study applied an established methodology for musculoskeletal model creation based on patient CT scans ^27,35,52,53^, including adjustment for spinal curvature and muscle measurements. However, the patient cohort and the healthy controls CT data were collected using different scanners and scan protocols. The resulting difference in CT scanner properties and image acquisition parameters (for example, slice-count, slice-thickness, dose optimization per patient habitus and reconstruction methods) ^54^ could affect model creation due to differences in segmentation and property estimation of the muscle and vertebrae affecting vertebral strength estimates. Although our group is investigating machine learning methods for image harmonization to account for such differences ^54^, this application is beyond the scope of this study. Spine loading was assessed for all individuals under standardized static poses. Static analysis underestimates loading from comparable dynamic scenarios by approximately 16% ^55^, which should not alter the overall findings. We note that using standardized poses does not capture any kinematic differences between individuals or groups, and the movement of patients with spine metastases remains unmeasured. Our study is a cross-sectional analysis study that does not directly address the risk or incidence of PVF. As noted, increased LSRs have been associated with both prevalent ^31–34^ and incident ^56^ OVF in older adults. An important limitation in the potential use of LSR in predicting vertebral fracture is that the modeled activities are normal daily tasks that are not expected to overload the spine to failure. Thus, while the LSR may be useful in predicting the fracture risk, they do not imply that these tasks will cause failure, nor do they suggest loading scenarios that might overload the spine. Despite these limitations, our study in vivo analyses of spine LSRs in patients with spine metastases is novel and highlights, for the first time, the effect of cancer on the difference in LSRs in patients from healthy normative values.

## Conclusion

This study uniquely determined vertebral LSRs in cancer patients and a healthy cohort, finding task-specific differences related to cancer, lesion type, and vertebral location. Overall, this initial assessment supports further examination of whether baseline LSR measurements are associated with incident PVF in patients with metastatic spine disease and, if so, what threshold values indicate risk. Our finding of lesion-mediated differences suggests that different thresholds for such measurements might need to be established based on vertebral region and metastatic lesion type. Uniquely, our finding that vertebrae with no observed metastatic lesions showed LSRs lower than the non-cancer cohort poses the question of whether these vertebrae should be considered “normal,” with important implications for patient management and risk reduction.

## Methods

Figure 5 presents a graphical summary of the protocol to create the musculoskeletal model and compute the vertebral load-strength ratio (LSR).

**Figure 5.**
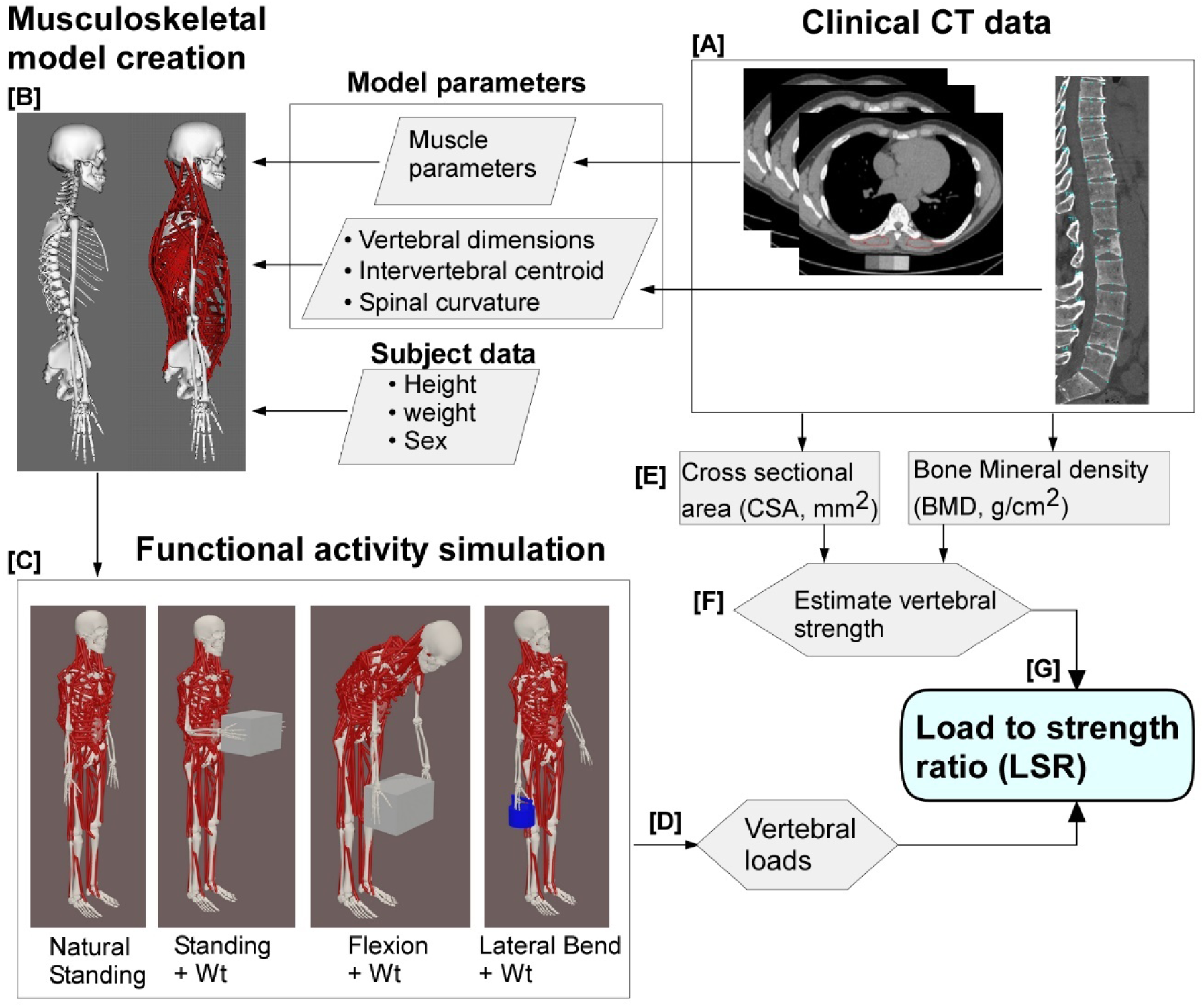
The workflow for establishing individualized cancer patient and FHS subjects’ musculoskeletal (MSK) models to estimate task-specific vertebral loading is as follows. The patient/ subject CT data is used to extract the subject-specific information [A: sex, height/ weight, muscle morphology, and spinal curvature, required to adjust our generic MSK model to the subject [B] ^35^. We then apply external loading and postures to simulate our selected sets of activities (Natural standing, standing with a weight of 10kg (Wt), Forward Flexion + Wt and Lateral Bending + Wt) (C). We use static optimization to predict muscle forces and calculate spinal joint loading using the joint analysis tool in OpenSim ^35^. The outcome of this analysis is individual vertebral loading [D]. From the CT data, and for each vertebral level, we estimate the integral bone mineral density (iBMD) and measure the vertebral cross, sectional area (CSA), [E], from which we used regression equation established in QCT-based finite-element analysis (FEA) ^63^ and validated in cadaveric cancer vertebrae ^36^, to predict vertebral strength from the CT data [F]. We then use the modeled vertebral loading and predicted strength to calculate the vertebral specific load-to-strength ratio (LSR) [G].

## Cancer cohort

The study cancer cohort comprised patients with metastatic spinal cancer who received radiation therapy at Dana Farber Cancer Institute or Brigham & Women’s Hospital between September 2020 and July 2023. All patients in this study had previously consented to the Broadband biorepository research project (MGB IRB 2016P001582). Study inclusion criteria were 1) the patient had histologically or cytologically documented stage IV bone metastases and radiographic (computed tomographic [CT] scan or bone scan) evidence of bone metastases and 2) Karnofsky Performance Status ^57^ >70. This criterion was selected to enhance the likelihood of patient participation and allow for the planned follow-up. Patients were excluded if they had: 1) Diseases of abnormal bone metabolism (including Paget disease, untreated hyperthyroidism, untreated hyperprolactinemia, untreated Cushing disease); or 2) If the spine region under consideration had previously undergone radiotherapy (< 6 months), surgery, or vertebral augmentation at the site of radiation or adjacent levels.

### 2.2 CT imaging data protocols

#### a. Cancer patient cohort

Cancer patients planned for radiotherapy were simulated for treatment at the radiation oncology Department, Brigham and Women’s Hospital by our study clinical senior attendings (TB, AS, and MAH, BWH Radiation Oncology, 17, 10 and 8 years of experience respectively), using the Siemens SOMATOM Confidence (Siemens Healthcare GmbH, Erlangen, Germany) or GE Lightspeed (General Electric Medical System, Waukesha, WI) CT scanners. Simulation scan parameters were: Tube voltage:120 (kVp); Tube current 240-300(mA); Field of view: A (16 cm) or B (Skin-to-Skin); Slice thickness: Siemens (For Stereotactic body radiotherapy (SBRT):0.5mm, All others: 1.5mm) GE Lightspeed (SBRT, All others: 1.25mm); in-Plane Pixel Size (mm), *A: 0.31*0.31, B: 0.70-0.98;* Gantry rotation: 1 second; Gating: None; Breath Hold: None.

#### b. Normative dataset: Framingham Heart Study (FHS)

To provide for comparison of LSRs from patients with spine metastases to LSRs from healthy adults, we drew on the same normative dataset from our prior analysis of LSRs in cadaveric metastatic spines ^35,36^. This normative dataset comprised a sample of men and women from the community-based Framingham Heart Study Multidetector CT Study ^58^, age- and sex-stratified to comprise equal numbers of women and men, ranging from 41 – 90 years of age ^59^. All participants in this study underwent abdominal and thoracic scans on a GE Discovery VCT 64-slice PET/CT scanner (GE Healthcare), with the following scan settings: a tube voltage of 120 kVp, tube current of 300/350 mA (≤ 220/> 220 lb body weight), and gantry rotation of 350 ms. These acquisitions typically included vertebral levels in the range of T4 – L4.

#### c. CT-based measurements and estimates of vertebral strength

Using a semi-automated image analysis program (Analyze 12.0, AnalyzeDirect, Inc., Overland Park, KS, U.S.), the mid-height of the vertebrae was identified on the CT mid-sagittal plane. The axial CT image corresponding to the identified vertebral mid-height was selected, and the vertebral body was segmented, including the proximal mid-pedicles. The segmentation was used to compute the vertebral cross-sectional area (CSA) and volumetric bone mineral density (BMD). Vertebral strength (Vs) was estimated in all subjects at all levels included in the CT scan using a previously developed regression equation ^35^,

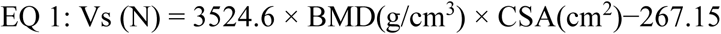

Trunk muscle size and positions were measured in all CT scans for use in creating musculoskeletal models according to previously described methods.

#### c. CT-based evaluations of metastatic lesions

All patient CT scans were reviewed by an expert neuroradiologist (DBH, more than 40 years of clinical experience) and spine biomechanics expert (RNA, 27 years of experience in vertebral imaging and biomechanics), with each vertebral level classified by agreement for the presence of metastatic lesions. The resulting classification categories of 1) no observed lesion (NOL), 2) Osteolytic, 3) Osteosclerotic, and 4) mixed were used for analysis.

#### d. Subject-specific musculoskeletal modeling

Using established methods, we created individualized musculoskeletal models for each cancer patient and FHS subject using existing generic full-body musculoskeletal models for men and women (Model version Fullbody_OS4.x_v2.0, available at https://simtk.org/projects/spine_ribcage). The models incorporated full thoracolumbar spines and included 620 musculotendon actuators, 78 rigid bodies, and 165 degrees of freedom ^27,52,53^. Models were adjusted to match patient height and weight, CT-based measurements of sagittal-plane spine morphometry and trunk muscle size and position, according to previous methods in other studies, including the normative dataset ^35^. Models were created, and model evaluations were performed in OpenSim version 4.3 ^60^ using custom scripting in Matlab (The Mathworks, Inc., Natick, MA, USA).

#### e. Evaluation of spine loading and LSR

Spine loading for patients and controls was assessed for four simulated static loading conditions previously analyzed in our cadaveric study ^36^. These include 1) neutral standing (NS); 2) standing with weight, wherein the elbows were flexed 90° and each hand held a weight of 5 kg (S+W); 3) forward flexion with weight, wherein a total of 60° of forward flexion was applied across the pelvis and spine, and each hand held a weight of 5 kg (F+W); and 4) lateral bending with weight, wherein a total of 20° of lateral bending to the right was applied to the spine, while the right hand held a weight of 5 kg weight in right hand (LB+W).

Static optimization analyses were performed in OpenSim for each subject and condition to evaluate muscle activations and forces required for the activity, and muscle force outcomes were subsequently used to calculate vertebral body compressive loading. These analyses minimized the sum of muscle activations cubed while determining muscle forces that would produce the required moments to balance all model joints in the applied posture. The resulting joint reaction forces, including the effect of muscle forces, were then determined for each vertebral joint and used to estimate the vertebral compressive loading at each vertebral level. Level-specific LSRs were defined as the vertebral compressive loading from the musculoskeletal model divided by the corresponding CT-based vertebral compressive strength for each patient and loading condition.

#### f. Data and variables

This study’s primary outcome variables of interest are the LSRs for each vertebral body from the T4 to L4 levels of spines from cancer patients and FHS cohorts. In order to put into context and highlight any observable difference in LSR between the groups (FHS and Patients), Standardized LSR was calculated for each vertebral level by referring to the LSR estimated in the corresponding vertebral levels in the normative FHS data in NS by the following formula:

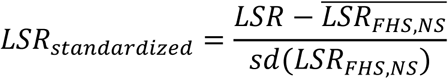

The standardization was done separately for males and females, and the raw LSR values were converted into standardized LSR, referring to the distribution obtained from the healthy subjects in NS. Body Mass Index was calculated using the standard formula ^61^. LSR outcomes, generated at each vertebral level, were categorized into one of three spinal regions for statistical analysis: Thoracic (T4-T10), Thoracolumbar (T11 – L1), and Lumbar (L2 – L4).

#### g. Statistical Analysis

The Statistical Software SAS® software (version 9.4 for Windows, SAS Institute, Cary, NC) was used to generate the descriptive statistics and to implement the models of interest. Appropriate descriptive statistics (mean and standard deviations for continuous variables; absolute and relative frequencies for nominal variables) were generated to describe the characteristics of the study participants (Age, Sex, Weight, Height, BMI) by group (FHS cohort vs patients with Metastatic Cancer). Similarly, the summary statistics for LSR were calculated separately for each task (NS, S+W, F+W, and LB+W). Statistical tests were used to check for any difference by a group for nominal (chi-square test) and continuous (Independent Samples t-test or, alternatively, the Mann-Whitney Test) variables.

Linear mixed models were used to estimate and test the differences in standardized LSRs for each task and sex separately between the FHS and type of lesion in Cancer Patients while accounting for the correlated nature of the data (i.e., LSR was calculated for different vertebral levels within the same individuals). The models were implemented to estimate the effects of Group (Healthy (FHS), NOL, Osteolytic, Osteosclerotic, Mixed), Age, Spinal Region, BMI, and their Interactions (Age × Group, Spinal Region × Group) on standardized LSR scores. The assumptions of the linear mixed models were verified, and the presence of outliers and influential observations were examined. Least Square Means differences are calculated as linear combinations from the model results to investigate differences by group within each spinal region, by sex and by task with a focus on comparing the lesion types (NOL, OL, OS, MX) with FHS cohort, and within Cancer cohort, comparing OL, OS, MX with NOL. The results of each model (coefficient estimate, standard error, and P value) are presented in tabular form, and the summary of differences in least-square means is presented in graphical form, created using the *ggplot* package in R ^62^. The alpha significance level was set at 0.05, and the p-values were adjusted using the False Discovery Rate approach ^63^.

## Data Availability

All data produced in the present study are available upon reasonable request to the authors

## Acknowledgments

The National Institute of Arthritis and Musculoskeletal and Skin Diseases supported the corresponding author, R. Alkalay, and the work of D. Anderson under its Research Project Grants (AR055582, R56AR075964, and AR075964). From the Framingham Heart Study of the National Heart Lung and Blood Institute of the National Institutes of Health and Boston University School of Medicine. This project has been funded in whole or in part with Federal funds from the National Heart, Lung, and Blood Institute, National Institutes of Health, Department of Health and Human Services, under Contract No. 75N92019D00031.

The authors would like to acknowledge the BROADBAND Research Project at the Brigham & Women’s Hospital Department of Radiation Oncology for providing regulatory and personnel support for this project. The BROADBAND Project was partly made possible by the generous donations of Stewart Clifford, Fredric Levin, and their families.

## Author Contributions

DEA, TB, DH and RNA conceived and designed the study. RNA, DEA, MK and drafted the manuscript. DK, PFD, HK, SK, SC, TB, AS, and MAH acquired and organized clinical data from patients. DEA, JJ, and BT performed musculoskeletal modeling and associated programming and analysis for patients and the normative database. JJ computed CT-based vertebral strength. DH and RNA performed the image-based metastatic bone lesions classification. MK performed statistical analysis. DEA, MK and RNA drafted and revised the manuscript. All authors have reviewed and approved the final manuscript.

## Supplemental Information

**Supplemental Table 1A:** Results of LMEMs comparing standardized LSRs for loading tasks by group and stratified by sex.

|  | NS |  |  |  | S + W |  |  |  |
| --- | --- | --- | --- | --- | --- | --- | --- | --- |
|  | Male |  | Female |  | Male |  | Female |  |
|  | B (SE) | P val. | B (SE) | P val. | B (SE) | P val. | B (SE) | P val. |
| **Intercept** | -0.014 (0.082) | 0.862 | -0.081 (0.066) | 0.250 | 4.521 (0.204) | <.001 | 4.206 (0.167) | <.001 |
| **Group\*** |  |  |  |  |  |  |  |  |
| MX | -0.817 (0.260) | 0.002 | -0.755 (0.193) | <.001 | -2.446 (0.739) | 0.002 | -2.485 (0.584) | <.001 |
| NOL | 0.020 (0.130) | 0.875 | -0.348 (0.130) | 0.030 | -0.449 (0.326) | 0.224 | -0.762 (0.338) | 0.065 |
| OL | 0.810 (0.174) | <.001 | -0.172 (0.196) | 0.433 | 0.969 (0.480) | 0.117 | -0.925 (0.600) | 0.181 |
| OS | -0.340 (0.157) | 0.056 | -0.670 (0.190) | 0.001 | -0.764 (0.424) | 0.082 | -1.035 (0.575) | 0.082 |
| **Age** | 0.008 (0.006) | 0.267 | 0.024 (0.005) | <.001 | 0.015 (0.014) | 0.407 | 0.069 (0.012) | <.001 |
| **BMI** | 0.058 (0.015) | <.001 | 0.060 (0.010) | <.001 | -0.034 (0.038) | 0.485 | 0.006 (0.026) | 0.804 |
| **Spinal region†** |  |  |  |  |  |  |  |  |
| THL | -0.009 (0.040) | 0.819 | -0.013 (0.033) | 0.781 | 1.248 (0.123) | <.001 | 1.245 (0.112) | <.001 |
| L | -0.005 (0.039) | 0.889 | 0.013 (0.032) | 0.798 | 0.107 (0.119) | 0.490 | 0.960 (0.110) | <.001 |
| **Interactions** |  |  |  |  |  |  |  |  |
| **Group×Age\*** |  |  |  |  |  |  |  |  |
| MX × Age | -0.042 (0.019) | 0.040 | -0.046 (0.014) | 0.006 | -0.070 (0.053) | 0.217 | -0.100 (0.043) | 0.039 |
| NOL × Age | -0.018 (0.010) | 0.142 | -0.029 (0.009) | 0.003 | -0.025 (0.026) | 0.376 | -0.070 (0.022) | 0.004 |
| OL × Age | -0.048 (0.012) | <.001 | -0.041 (0.012) | 0.001 | -0.066 (0.033) | 0.045 | -0.138 (0.035) | <.001 |
| OS × Age | -0.026 (0.012) | 0.079 | -0.026 (0.011) | 0.079 | -0.052 (0.032) | 0.167 | -0.023 (0.032) | 0.458 |
| **Group×Spinal Region\*†** |  |  |  |  |  |  |  |  |
| MX × THL | -0.043 (0.245) | 0.955 | -0.046 (0.191) | 0.955 | -0.757 (0.751) | 0.743 | -0.581 (0.650) | 0.743 |
| MX × L | -0.151 (0.262) | 0.564 | -0.129 (0.199) | 0.564 | -0.591 (0.805) | 0.564 | -0.545 (0.679) | 0.564 |
| NOL × THL | 0.436 (0.070) | <.001 | 0.150 (0.070) | 0.042 | 0.805 (0.212) | <.001 | -0.089 (0.238) | 0.710 |
| NOL × L | 0.397 (0.073) | <.001 | 0.299 (0.073) | <.001 | 0.928 (0.225) | <.001 | 0.230 (0.249) | 0.355 |
| OL × THL | 0.136 (0.167) | 0.997 | 0.014 (0.212) | 0.997 | 0.211 (0.512) | 0.997 | 0.375 (0.724) | 0.997 |
| OK × L | -0.648 (0.182) | 0.001 | -0.109 (0.276) | 0.792 | -1.275 (0.559) | 0.060 | 0.053 (0.940) | 0.955 |
| OS × THL | 0.429 (0.145) | 0.008 | 0.105 (0.191) | 0.602 | 0.309 (0.445) | 0.602 | -1.233 (0.650) | 0.116 |
| OS × L | 0.414 (0.141) | 0.014 | 0.479 (0.180) | 0.021 | 0.336 (0.434) | 0.584 | -0.233 (0.612) | 0.704 |

Reference category: \*: Group (FHS); † Spinal region (Thoracic); \*\*: variables are centered by their average; B: Regression Coefficient; SE: Standard Error; NOL: no CT-observable lesions; OL: Osteolytic; OS: Osteosclerotic; NS: neutral standing; S+W: standing with weight; P values are adjusted for multiple testing by the FDR correction.

**Supplemental Table 1B:** Results of LMEMs comparing standardized LSRs for loading tasks by group and stratified by sex.

|  | F + W |  |  |  | LB + W |  |  |  |
| --- | --- | --- | --- | --- | --- | --- | --- | --- |
|  | Male |  | Female |  | Male |  | Female |  |
|  | B (SE) | P val. | B (SE) | P val. | B (SE) | P val. | B (SE) | P val. |
| **Intercept** | 4.033 (0.222) | <.001 | 3.412 (0.173) | <.001 | 2.274 (0.125) | <.001 | 1.936 (0.094) | <.001 |
| **Group\*** |  |  |  |  |  |  |  |  |
| MX | -2.154 (0.770) | 0.006 | -1.539 (0.574) | 0.007 | -1.611 (0.393) | <.001 | -1.391 (0.288) | <.001 |
| NOL | -0.597 (0.352) | 0.144 | -0.155 (0.347) | 0.750 | -0.383 (0.197) | 0.105 | -0.700 (0.189) | 0.002 |
| OL | 0.752 (0.504) | 0.181 | -0.278 (0.588) | 0.636 | 0.769 (0.264) | 0.014 | -0.492 (0.292) | 0.181 |
| OS | -0.944 (0.448) | 0.056 | -0.640 (0.565) | 0.257 | -1.038 (0.239) | <.001 | -1.183 (0.283) | <.001 |
| **Age\*\*** | 0.000 (0.016) | 0.978 | 0.068 (0.012) | <.001 | 0.005 (0.009) | 0.606 | 0.041 (0.007) | <.001 |
| **BMI\*\*** | 0.025 (0.041) | 0.612 | 0.088 (0.027) | 0.002 | 0.031 (0.023) | 0.298 | 0.058 (0.015) | <.001 |
| **Spinal region†** |  |  |  |  |  |  |  |  |
| THL | 3.143 (0.126) | <.001 | 2.800 (0.108) | <.001 | -0.485 (0.060) | <.001 | -0.464 (0.050) | <.001 |
| L | 4.319 (0.121) | <.001 | 4.657 (0.105) | <.001 | -0.249 (0.058) | <.001 | -0.128 (0.049) | 0.015 |
| **Interactions** |  |  |  |  |  |  |  |  |
| **Group×Age\*** |  |  |  |  |  |  |  |  |
| MX × Age | -0.017 (0.055) | 0.760 | -0.101 (0.042) | 0.039 | -0.054 (0.029) | 0.081 | -0.066 (0.021) | 0.006 |
| NOL × Age | -0.010 (0.028) | 0.732 | -0.069 (0.023) | 0.005 | -0.020 (0.016) | 0.275 | -0.047 (0.013) | 0.001 |
| OL × Age | -0.094 (0.035) | 0.008 | -0.110 (0.035) | 0.003 | -0.058 (0.019) | 0.003 | -0.066 (0.018) | 0.001 |
| OS × Age | -0.052 (0.034) | 0.167 | -0.042 (0.032) | 0.216 | -0.034 (0.018) | 0.129 | -0.036 (0.016) | 0.079 |
| **Group×Spinal Region\*†** |  |  |  |  |  |  |  |  |
| MX × THL | -1.733 (0.764) | 0.094 | -1.873 (0.621) | 0.021 | -0.021 (0.369) | 0.955 | 0.088 (0.292) | 0.955 |
| MX × L | -2.373 (0.819) | 0.015 | -2.855 (0.648) | <.001 | -0.341 (0.394) | 0.564 | -0.215 (0.305) | 0.564 |
| NOL × THL | 0.365 (0.216) | 0.105 | -0.871 (0.227) | <.001 | 0.494 (0.104) | <.001 | 0.253 (0.107) | 0.029 |
| NOL × L | 0.720 (0.229) | 0.003 | -0.821 (0.238) | 0.001 | 0.339 (0.110) | 0.003 | 0.252 (0.112) | 0.028 |
| OL × THL | 0.040 (0.521) | 0.997 | -0.379 (0.691) | 0.997 | -0.077 (0.251) | 0.997 | 0.001 (0.325) | 0.997 |
| OK × L | 0.390 (0.569) | 0.703 | -0.719 (0.897) | 0.703 | -1.299 (0.274) | <.001 | -0.267 (0.423) | 0.703 |
| OS × THL | -0.469 (0.453) | 0.482 | -2.238 (0.621) | 0.003 | 0.721 (0.218) | 0.004 | 0.152 (0.292) | 0.602 |
| OS × L | -0.246 (0.441) | 0.659 | -1.887 (0.585) | 0.010 | 0.544 (0.213) | 0.021 | 0.593 (0.275) | 0.050 |

Reference category: \*: Group (FHS); † Spinal region (Thoracic); \*\*: variables are centered by their average; B: Regression Coefficient; SE: Standard Error; NOL: no CT-observable lesions; OL: Osteolytic; OS: Osteosclerotic; F+W: forward flexion with weight; LB+W: lateral bending with weight; P values are adjusted for multiple testing by the FDR correction.

